# Accessibility and perceived quality of GP care for socioeconomically disadvantaged patients: a qualitative patient-centered interview study

**DOI:** 10.1101/2025.02.27.25322989

**Authors:** Hannah A. Bijl, Dionne N. Zwinkels, Reinier C.A. van Linschoten

## Abstract

**Introduction:** Patients with lower socioeconomic status (SES) in the Netherlands may face significant barriers to accessing and receiving general practitioner (GP) care. Factors such as lower health literacy, economic disadvantage, and cultural diversity contribute to these challenges, often leading to lower satisfaction and reduced access. This study investigates how patients with low SES perceive the accessibility and quality of GP care, with the goal of identifying ways to improve care for this vulnerable group.

**Methods:** This qualitative study used in-depth semi-structured interviews to explore how patients with low SES experience the accessibility and perceived quality of GP care. Data were analyzed using thematic analysis.

**Results:** A total of 16 patients were interviewed. Key themes emerged regarding accessibility and perceived quality of care. Patients highly valued quick access to appointments and multiple contact options. Digital healthcare tools were appreciated for their convenience, especially among younger and more educated patients. However, face-to-face interactions were universally preferred for serious health concerns. Effective communication and empathic care turned out to be crucial factors in shaping patient satisfaction. Lastly, the importance of continuity of care was emphasized, especially by patients with chronic conditions, who valued building strong rapports with their GP.

**Conclusion:** Our study found that quick access, multiple contact options, and clear communication are key drivers of patient satisfaction with GP care for patients with low SES. Emphasizing communication quality and empathetic care can significantly improve the perceived quality and accessibility of healthcare for vulnerable populations.

## Introduction

General practitioners (GPs) serve as the primary point of access to healthcare in the Netherlands. This system aims to ensure that patients receive timely and appropriate care. However, the increasing demand for GP services is placing significant pressure on the accessibility of GP care.(1) Research showed that the number of consultations and home visits in 2024 surpassed those of the previous year, and the waiting list for GPs has increased compared to previous years.(2) To ensure accessibility, GP services have to be reorganized.

Socioeconomic status (SES) plays a crucial role in the accessibility and perceived quality of GP care. SES is generally determined by income, educational level and employment status.(3) Patients with lower SES often face compounding barriers to accessing healthcare, including financial constraints, lower health literacy, and limited availability of services in their neighborhoods.(4-6) Studies have demonstrated a strong association between neighborhood socioeconomic disadvantage and access to healthcare, highlighting these compounding barriers faced by patients living in socioeconomically deprived areas.(7) This is particularly relevant in diverse urban environments, where populations tend to have lower SES and higher rates of chronic disease, often resulting in more GP appointments.(8-10) These factors, together with reduced levels of health literacy and cultural diversity, can create considerable obstacles to accessing GP care.

In addition to accessibility, SES also impacts the perceived quality of care. Studies have shown that patients with lower SES report lower satisfaction levels compared to patients with higher SES.(11) This difference in satisfaction is observed across various healthcare settings.(12) Perceptions of quality and access are shaped by structural factors, such as appointment availability and waiting times, as well as interpersonal factors, including communication with healthcare providers and trust in the care received.(11, 13-15) Perceived quality of care is closely linked to patient satisfaction, adherence to treatment plans, and health outcomes.(16) However, SES significantly influences these perceptions, with patients with lower SES often reporting lower satisfaction and facing more barriers to accessing high-quality care. Investigating how patients with low SES perceive the accessibility and quality of primary healthcare is essential for addressing health inequalities and improving health outcomes.

One promising and rapidly emerging solution to alleviate the increasing pressure on GPs and improve accessibility is the digitalization of healthcare.(17-19) Examples include allowing patients to schedule an appointment online or send electronic consultations (e-consults) where they receive a prompt and direct response to brief inquiries.(20) These digital solutions not only aim to support the GP by managing patient flow more efficiently, but also empower patients by offering flexible and immediate access to care. Despite the potential benefits, equitable access to digital healthcare remains a concern, particularly for patients with lower SES. These patients often have reduced digital health literacy, which is defined as the ability to find and use health information from digital sources.(21) Patients with lower digital literacy are less likely to use digital healthcare tools, such as video consultations, which limits their ability to benefit from these innovations.(22, 23) Because SES is an important determinant of digital health literacy, the increasing use of digital healthcare leads to concerns regarding the accessibility and perceived quality of GP care for patients with a lower SES.(24, 25)

Previous research has explored the relationship between SES and patient satisfaction, but there is little in-depth information about how patients from low SES backgrounds experience the accessibility and quality of care provided by their GPs and how care for these patients may be improved. We aimed to study how patients with low SES backgrounds experience accessibility and quality of GP care in the Netherlands, and how the quality and accessibility of GP care for these vulnerable groups can be improved.

## Methods

This study had a qualitative research design, using in-depth semi-structured interviews to investigate the experiences of patients with low SES with the accessibility and perceived quality of care of their GP. The research approach in this study was phenomenological. We have chosen this approach to allow for an in-depth understanding of personal experiences, perceived barriers, and potential recommendations to improve accessibility and quality of care for this group of patients.

The following main question guided our investigation: How do patients with a low SES perceive the accessibility and quality of healthcare services provided by their GP? Sub-questions we aimed to address were:

- What barriers do they encounter when accessing general practitioner services?
- What expectations do these patients have of their GP and how do these expectations influence their experience with care?
- How does the timeliness and availability of appointments impact the perceived quality of care?
- What role does the use of digital healthcare play in shaping the perceived quality of care for these patients?

### Researcher characteristics and reflexivity

The first author conducted all interviews. She was a medical student at Erasmus University Rotterdam who started her studies in 2018 and recently finished her clinical rotations, which provides a comprehensive understanding of the healthcare system and the challenges faced by both patients and practitioners. The author had limited interview experience and was not known to the participants of this research before undertaking the study.

### Context

Patients were sampled from a general practice in Rotterdam-Feijenoord. Rotterdam is one of the most socioeconomically heterogeneous cities in the Netherlands, characterized by a wide range of SES-WOA (wealth, educational attainment, and labor market participation) scores. This heterogeneity is exemplified by the stark contrast between affluent neighborhoods like Hillegersberg and Kralingen and economically disadvantaged areas such as Rotterdam-Feijenoord. Rotterdam-Feijenoord has a population of approximately 7,700 residents, of whom 80% have a non-Dutch background. About three-quarters of the households fall into the lowest income class, a proportion considerably higher than the average in Rotterdam.(26, 27) By situating this study in Rotterdam-Feijenoord, we aimed to capture the challenges and experiences of patients from one of the city’s most socioeconomically disadvantaged and culturally diverse areas.

### Participant selection and sampling strategy

We aimed to include adult patients with low SES from a single general practice in Rotterdam-Feijenoord. This was a relatively new practice which opened in January 2024. Patients were included if they had an educational level below MBO (secondary vocational education). Participants were recruited by purposive sampling based on a sampling matrix. This sampling matrix included employment status, migration background, language barrier and preferred way of contacting the GP. The matrix aimed to capture a diverse group of patients, to enhance the relevance and transferability of our findings to other populations. Patients were selected by the treating GPs and then invited and asked for informed consent to participate in the study by the researcher. Interviews were conducted until data saturation was achieved, meaning no new themes or insights were emerging from the data.

### Ethical approval

This study was conducted as a master thesis in collaboration with Jans Huisartsen and the department of General Practice of Erasmus MC. A waiver for the Medical Research Involving Human Subjects Act was provided by the institutional review board of the Erasmus MC. All patients were informed about the study’s purpose, procedures and their rights as participants. Participation was voluntary, and patients were assured they could withdraw from the study at any time without any consequences. Informed consent was obtained from all participants prior to their involvement in the study.

### Data collection and processing

Data were collected through semi-structured interviews conducted between August and September 2024. Interviews lasted around 20 minutes and were conducted face-to-face at the general practice. The interview guide included open-ended questions on themes of experience with accessibility and perceived quality of care of the general practice, barriers encountered when making an appointment and possible recommendations to improve the accessibility or quality of care (Supplementary File 1). Prompting and probing were used as techniques to encourage participants to elaborate on their responses and to gain deeper insights into their experiences. This involved using follow-up questions such as “Can you tell me more about that?” or “What do you mean by that?” as well as requesting clarification or examples when participants provided brief or unclear answers. All interviews were recorded with MS Teams and transcribed verbatim after. Data were anonymized by assigning numbers per patient. For each interview date, time and place of interview, patient code and name of interviewer and exact duration of the interview were documented. In addition, each participant’s age, education level, employment status, country of birth, country of birth of parents, and presence/absence of chronic illness were recorded before starting the interview.

### Data analysis

The transcripts were analyzed using thematic analysis. MAXQDA software was used to facilitate the coding process. Initially, the first author conducted the coding independently, after which another researcher coded a subset of transcripts. Based on this, the coding tree was further refined. Afterwards, the first author recoded all transcripts using the updated coding tree. Open coding was first performed to identify significant statements and phrases relevant to the research questions, followed by axial coding to organize these into broader (sub)themes. An overview of the codes used can be found in Supplementary File 2.

## Results

As seen in Figure 1, a total of 19 patients were approached to participate. Of these, two patients failed to attend their scheduled interview appointments. The remaining 17 patients were interviewed. However, during the course of the interview it was determined that one participant did not meet the low SES criterion. This participant was therefore excluded from the final analysis. Thus, the final sample consisted of 16 patients who all met the inclusion criteria and completed the interview. Data saturation was reached during the interviews, as no new themes emerged after the analysis of the final participants’ responses.

**Figure 1.**
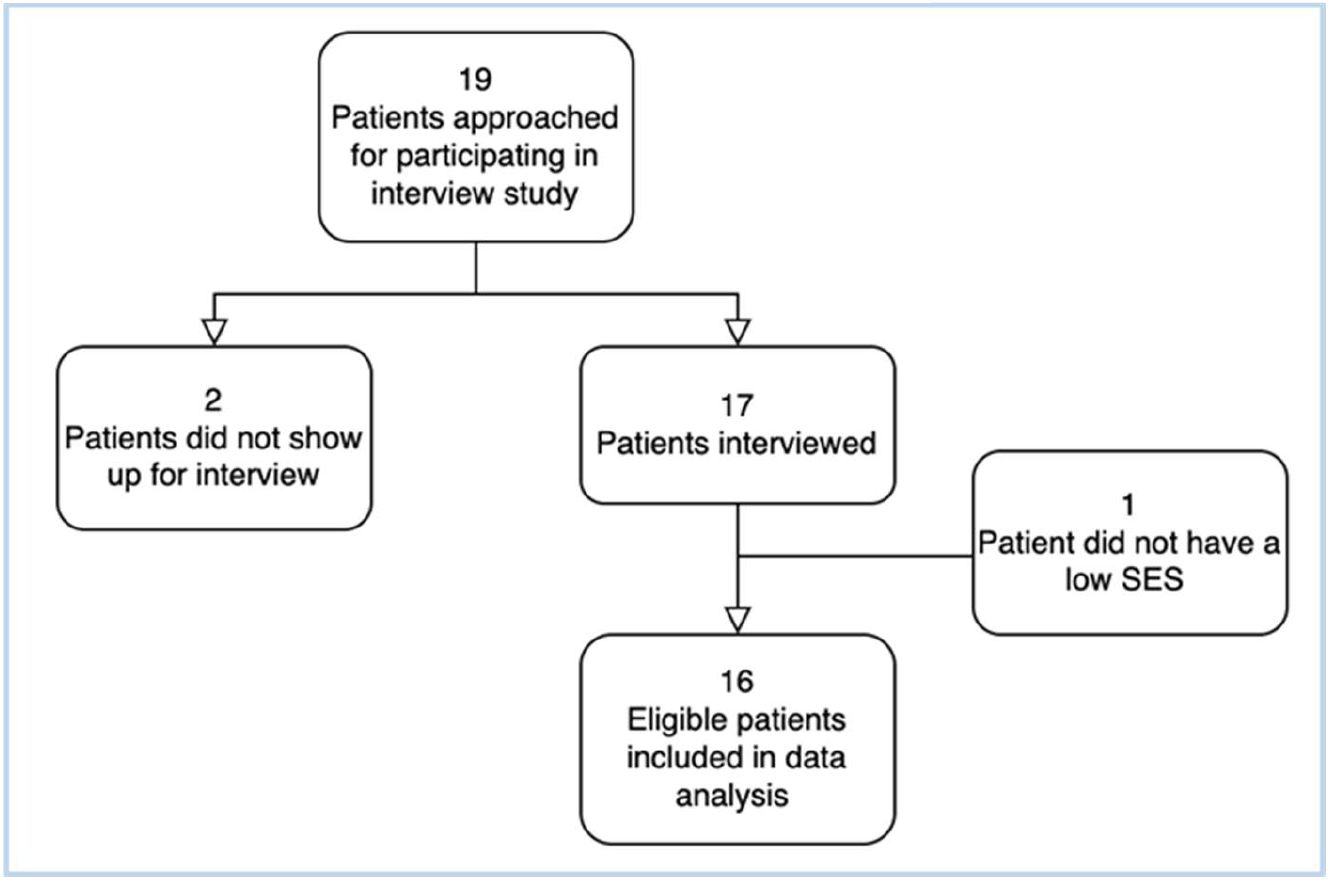
Flowchart of the participants included.

Of all participants included, the majority were female (Table 1). More than half of the patients reported having a chronic illness. The educational levels of the participants varied, ranging from no formal education to vocational education. Most participants, as well as their parents, were of Dutch origin, although there were several participants with migrant backgrounds from countries such as Albania, Turkey and Suriname.

**Table 1.** Patient characteristics (n = 16)

|  |  | *n* | (%) |
| --- | --- | --- | --- |
| **Age range (years)** | 24-69 |  |  |
| **Sex** |  |  |  |
|  | Male | 5 | (31) |
|  | Female | 11 | (69) |
| **Educational level** |  |  |  |
|  | No school or education completed | 5 | (31) |
|  | Pre-vocational education | 4 | (25) |
|  | Secondary vocational education | 7 | (44) |
| **Country of birth** |  |  |  |
|  | Netherlands | 15 | (94) |
|  | Dutch Caribbean | 1 | (6) |
| **Country of birth mother** |  |  |  |
|  | Netherlands | 11 | (69) |
|  | Europe, other | 2 | (13) |
|  | Turkey | 1 | (6) |
|  | Dutch Caribbean | 1 | (6) |
|  | Suriname | 1 | (6) |
| **Country of birth father** |  |  |  |
|  | Netherlands | 10 | (63) |
|  | Europe, other | 2 | (13) |
|  | Turkey | 1 | (6) |
|  | Dutch Caribbean | 1 | (6) |
|  | Suriname | 1 | (6) |
|  | Asia | 1 | (6) |
| **Employment status** |  |  |  |
|  | Employed | 10 | (63) |
|  | Unemployed | 6 | (37) |
| **Chronic illness** |  |  |  |
|  | Yes | 9 | (56) |
|  | No | 7 | (44) |

## Accessibility of GP care

We found that nearly all participants expressed high levels of satisfaction with their practice’s accessibility. This was primarily linked to the speed with which patients were attended to. Participants reported that the practice was always reachable via telephone and via an online application, that they had minimal waiting times for their calls to be answered, and that they were able to secure doctor’s appointments without significant delays.

Patients expressed varying preferences in how they preferred to make appointments, with some preferring to physically visit the practice, while others opted to call or use the online application. One participant preferred to ‘just walk in’ and make an appointment in person, especially given their proximity to the practice: “*I live nearby, so I usually just walk in and make the appointment at the desk*.” This was seen as convenient for those who had additional questions. Another patient preferred telephone contact, stating that it allowed for immediate responses: *“When I call, I get an answer straight away. It’s quicker than using the app, where it takes longer to explain what you need*.*”* Several participants did mention using the app but noted that they often still chose to call for urgent matters.

### Barriers encountered when accessing GP services

Key barriers – mainly mentioned in patients’ experiences with their previous practices and not in this current practice – included long waiting times for appointments and difficulties in contacting the practice. Patients reported experiences of having to wait weeks for an appointment, leading to feelings of dissatisfaction and concerns about their health needs being unmet. A participant reflected, *“At my previous GP, I had to wait three or four weeks for an appointment. Or you could go to a walk-in consultation, but that was always completely full*.*”* Furthermore, strict triage protocols were noted as barriers to accessing care. Some patients expressed feeling dismissed or not taken seriously when they requested an appointment, stating, *“They go really deep into their questions, and when you’re not feeling well, it’s really frustrating to answer so many questions*.*”*

### Impact of timeliness and availability of appointments

The timeliness and availability of appointments influenced patients’ perceived quality of care. Quick access to appointments was often mentioned in conjunction with higher satisfaction levels. Many participants noted that being able to secure an appointment within a few days improved their overall perception of the practice. For instance, one patient stated, *“I can usually get an appointment within one or two days, and that feels good*.*”* Conversely, long wait times contributed to a sense of neglect and dissatisfaction. Patients often expressed frustration over delays in obtaining repeat prescriptions, stating, *“If I call for a repeat prescription, it takes too long to get a response*.*”*

### Role of digital healthcare

The integration of digital tools, such as apps and online consultations, has become increasingly important for patients, particularly considering the evolving technological landscape. It was primarily the younger generation of participants who expressed enthusiasm for the use of digital tools, with one participant remarking, *“This is the new era,”* acknowledging the convenience they offer for non-critical issues. Many younger patients appreciated the efficiency of the practice’s app, highlighting features such as quick response times: *“They respond within ten minutes, and I can book appointments or ask questions through the app. It’s really convenient*.*”* However, concerns about the limitations of digital consultations were also expressed. One participant emphasized, *“You can’t make a diagnosis over the phone or from photos. A picture doesn’t show what’s really happening beneath the surface*.*”*

While younger patients or those more familiar with technology found the app beneficial, the older generation remained skeptical about its accuracy and thoroughness. Although many reported receiving timely responses, some noted variability in response times, stating, *“Sometimes they say it’ll be thirty minutes, but it can take up to two or three hours, depending on the question*.*”* Others also mentioned limitations, for example a lack of functions such as asking for repeat prescriptions and unclear response times. Despite these occasional delays, many participants found the app useful for non-urgent matters.

Secondly, participants with a higher educational background, particularly those at MBO-level (secondary vocational education), appeared more receptive to digital communication tools compared to patients with lower or no educational backgrounds. This group often emphasized the practicality and efficiency of these tools in managing routine healthcare interactions. One participant noted, *“Using the app saves time and avoids unnecessary visits*.*”*

Besides these advantages, a common sentiment among participants was a preference for face-to-face interactions. Participants expressed a clear preference for in-person consultations, citing the ability for physicians to conduct physical examinations as a primary advantage. One participant remarked, *“I might be considered old-fashioned, but I prefer face-to-face consultations. I believe that when I’m physically present with the doctor, they can quickly check me if necessary, which is not possible over the phone*.*”* While some acknowledged the convenience of telephone consultations, particularly for simple matters like receiving test results, the general consensus was that for more serious health concerns requiring examination, in-person visits were preferred. *“For an examination, phone consultations don’t suffice; the doctor can’t see what’s going on”*, one participant noted. Nearly all patients emphasized the importance of personal contact in healthcare, with one respondent stating, *“I prefer to speak directly with someone; it feels more personal*.*”*

## Perceived quality of GP care

Participants articulated distinct perceptions regarding the perceived quality of healthcare services provided by their GPs. Their experiences reveal a nuanced understanding of the factors influencing their (dis)satisfaction with GP care.

### Expectations of GPs

One of the primary expectations shared by patients was the need for clear communication and understanding. Many emphasized the importance of receiving straightforward, comprehensible information. As one participant noted, *“I just want to understand what is being said to me. Clear language is crucial*.*”* Another key expectation was the establishment of a strong rapport with their GP, which was particularly important for patients with chronic conditions. Many of these participants emphasized the value of seeing the same GP each visit to build a strong rapport over time. One individual remarked, *“The GP is a crucial person in my life; I need to feel comfortable sharing my concerns and not having to tell my story over and over again*.*”*

The desire for active participation in decision-making also emerged as a recurring theme. Many patients expressed that they wanted their GPs to involve them in choosing their treatment options. One participant explained: *“I want my GP to ask what I think about my treatment options. It’s my health, after all*.*”* Lastly, accessibility and availability of care were frequently mentioned as critical factors. Participants stressed the importance of being able to access timely care, particularly when they were unwell. As one individual remarked, *“When I feel unwell, I need to see a doctor quickly. Long waits are not an option for me*.*”* This highlights the urgency often felt by patients, especially those with complex health needs, to receive prompt attention from their GP.

### Satisfaction with GP services

Satisfaction levels among patients were closely linked to the fulfillment of their expectations, with several key factors contributing to their overall satisfaction. One of the most important factors - which was notably not mentioned in the expectations of GP care - was the quality of care received. Patients expressed high levels of satisfaction when their health issues were effectively addressed. As one individual remarked, *“Since I’ve been registered here, I’ve received excellent care. My doctor is proactive in finding solutions*.*”* Continuity of care also emerged as a vital aspect of patient satisfaction. Many participants expressed a strong preference for seeing a consistent GP who was familiar with their medical history. One patient shared, *“Having the same doctor who knows my history makes it easier to discuss my health*.*”*

The emotional aspects of care, particularly empathy and personal attention, were highlighted as key contributors to satisfaction. One participant noted, *“When my doctor listens and shows genuine concern, it makes a world of difference. I leave feeling understood*.*”* Last, the environment of the practice also played a role in shaping patient satisfaction. Some participants commented on the importance of a welcoming and comfortable setting. One individual said, *“I feel at ease in this clean and modern practice. It’s important to me*.*”*

### Sources of dissatisfaction

Conversely, dissatisfaction among patients was often due to unmet expectations. One major source of frustration was inadequate communication. Many patients reported feeling ignored or overlooked when their questions were not clearly answered. One participant stated, *“I often feel like my questions are ignored. It leaves me feeling neglected*.*”*

Inconsistent care also emerged as an issue, particularly when patients did not have a designated GP. One individual remarked, *“Seeing a different doctor each time makes it hard to build a relationship. I don’t want to repeat my story every visit*.*”* This lack of continuity made it difficult for patients to develop trust. Unfriendly interactions with healthcare providers also fueled dissatisfaction. Instances of perceived rudeness or dismissive behavior were noted by several patients. One participant stated, *“If a doctor is dismissive, it makes me less likely to seek help again*.*”*

Finally, insufficient problem-solving was a recurring issue. Patients expressed frustration when their concerns were not adequately addressed. One participant shared, *“If I leave without a clear solution, I feel hopeless about my health*.*”* This lack of effective resolution led to feelings of helplessness and sometimes disengagement from seeking further care.

## Discussion

We found that patient satisfaction with GP accessibility was primarily influenced by quick access to appointments and multiple contact options (phone, in-person, app). While digital tools offered convenience, all patients expressed a preference for face-to-face interactions for more serious concerns. For patients with low SES, satisfaction with GP care was closely linked to effective communication, rapport-building, and empathetic care, while dissatisfaction often stemmed from communication issues, inconsistent care, and inadequate problem-solving.

In our study, we found that most participants reported high levels of satisfaction with the accessibility and quality of care provided. A likely explanation for this is that the practice from which we sampled our participants is relatively new and therefore not yet experiencing high patient demand. This lower patient load allows for quicker access to care, which emerged as a key factor in patient satisfaction. This finding aligns with previous research, which emphasizes that waiting times are a critical determinant of patient satisfaction in different health care settings.(28, 29)

Another finding from our study was the strong preference for seeing a regular GP, mainly among patients with chronic diseases. These patients emphasized the importance of continuity of care and the value of not having to repeatedly explain their medical history. The desire for a consistent healthcare provider among chronically ill patients underscores the importance of relationship-building in healthcare. Seeing a regular GP who is familiar with a patient’s medical history can provide more personalized care, potentially leading to better health outcomes and increased patient satisfaction.

Analysis of patient preferences regarding methods of contact with the practice revealed heterogeneous responses. Notably, the data suggest that age was the primary determinant of which communication channels were preferred. We observed that younger patients predominantly used the online application when contacting the practice, while the older patients tended to prefer calling or visiting the practice in person to schedule an appointment or ask their question. Additionally, participants with a higher educational background also showed a strong preference for using the app, citing its efficiency and ease of use. However, for more urgent matters, nearly all patients called the practice to ensure a direct response.

Our findings highlight a critical balance that must be struck in healthcare: reducing workload through digital tools while still ensuring a patient-friendly experience. Although digital platforms may potentially streamline processes and reduce work pressure for staff, they may not meet the needs of all patients. Many patients value the direct human interaction that traditional methods, like phone calls or in-person visits, provide. This reliance on digital access, while possibly beneficial in efficiency, raises concerns about the quality of care and the personal connection that many patients still seek. It is essential to consider whether digital solutions are truly the best approach if patients do not necessarily prefer them. While digital tools offer convenience, they should not replace the personal touch that many patients value in their healthcare experience. Maintaining multiple communication channels to accommodate varying patient preferences is crucial. This approach allows practices to meet the push for digital access without excluding those who value a more personal touch in their healthcare experience. Ultimately, the goal should be to enhance efficiency without compromising the quality of care and patient satisfaction.

The findings also suggest that solving medical issues is not the most critical determinant of patient satisfaction and experienced quality of care for patients with low SES. Rather, the quality of communication between patients and healthcare providers appears to be the decisive factor. Patients emphasized that clear and empathetic communication, coupled with the ability of providers to build strong rapports, played a more significant role in shaping their overall satisfaction than the resolution of their health problems.

In line with our findings, previous studies have highlighted that communication barriers, particularly in culturally and linguistically diverse populations, play a key role in shaping patient satisfaction.(30) Effective communication, trust-building, and empathy from healthcare providers are central to how patients perceived the quality of care. This aligns with existing literature that emphasizes the importance of interpersonal skills in healthcare settings, especially for vulnerable populations, such as those with low socioeconomic status.(31-34) Patients with low SES, who may already face barriers like limited health literacy, economic constraints, or unfamiliarity with healthcare systems, rely heavily on clear communication. Research suggests that healthcare providers need to adapt their communication style, as low SES patients often take a more passive role in consultations and engage in less relational talk.(35)

To address these communication disparities, healthcare providers may need to become more aware of differences in patient communication styles.(34) Strategies such as using a question prompt list - a structured list of questions related to illness, treatment and/or support - may help patients with lower SES raise concerns during their consultations. This can potentially reduce inequalities in communication and health outcomes.(35) Additionally, reducing the time pressures on healthcare professionals, through measures such as longer consultations or support from care coordinators, could allow for more meaningful interactions, ultimately improving patient satisfaction and health outcomes.(36)

### Strength and limitations

One of the strengths of our study is that we managed to conduct interviews until data saturation was achieved, despite the challenges of recruiting patients with a low SES who are generally hard to reach. Moreover, we managed to include patients from diverse educational levels.

A limitation of this study is the selection bias from recruiting participants exclusively from a single practice. Since participants were selected based on prior visits to this practice, the sample may not reflect patients who face difficulties accessing care or those who don’t visit the practice often. Additionally, results may be influenced by factors unique to this specific practice such as appointment availability, rather than offering insights applicable to other practices. This limits the generalizability of our findings to broader patient populations. Future research should aim to include patients from multiple practices and actively seek out those who struggle to secure appointments to capture a fuller picture of access barriers, especially among vulnerable groups.

A second limitation is that, despite using a comprehensive sampling matrix, we likely missed certain patient types. The matrix consisted of 24 possible combinations based on employment status, migration background, language barriers, and mode of contact (phone, digital, walk-in). We only interviewed 16 patients when data saturation was achieved. This may limit the generalizability of our findings, as the experiences and perceptions of these unrepresented patient types might differ from those included in the study.

Another limitation of this study is its focus on English and Dutch-speaking participants, excluding non-fluent speakers - particularly migrants - who may face additional healthcare barriers. This exclusion may underrepresent vulnerable groups affected by language challenges. Future studies could address this by using multilingual interviewers or translation tools to capture a wider range of perspectives.

Additionally, while the sample included diverse backgrounds, it may not fully reflect the region’s demographic diversity, which includes patients with origins in Morocco, Turkey, and Indonesia.(37) This underrepresentation could also affect the generalizability of our findings. Expanding recruitment to include these groups in future research would enhance the study’s generalizability.

### Future perspectives

Looking ahead, it is crucial to explore how digital healthcare solutions can be tailored to meet the diverse needs of all patient groups, particularly those with low SES. Future research should investigate the effectiveness of hybrid models that combine digital and traditional methods to ensure inclusivity and high-quality care. Moreover, studies could focus on developing and testing interventions aimed at improving digital health literacy among populations with lower SES, ensuring they can benefit from technological advancements in healthcare.

In practice, healthcare providers should keep focusing on training in communication skills and cultural competence to better serve diverse patient populations. Implementing policies that allow for longer consultation times and the use of care coordinators could potentially help reduce the barriers faced by patients with low SES, leading to better health outcomes and higher patient satisfaction. By addressing these areas, we can move towards a more equitable healthcare system that employs digital innovations while maintaining the personal touch that is proven to be essential for patient-centered care.

## Conclusion

Our study highlights that patient satisfaction with GP accessibility is driven by quick access to appointments and the availability of multiple contact options, with a clear preference for in-person consultations for serious health concerns. Digital tools are appreciated, particularly by younger and the higher educated patients, but face-to-face interactions remain crucial for complex issues. For patients with low SES, communication quality and empathetic care are key factors influencing their perceived quality of care. By recognizing that communication, rather than the resolution of medical issues, is central to patient satisfaction, healthcare systems can better address the needs of patients with low SES.

## Supporting information

Supplementary file 1. Interview guide

Supplementary file 2. Interview excerpts

## Data Availability

All data produced in the present study are available upon reasonable request to the authors through DataverseNL.

https://doi.org/10.34894/9CA1W1

