## Supplementary file 1. Interview guide for "Accessibility and perceived quality of GP care for socioeconomically disadvantaged patients: a qualitative patient-centered interview study"

### Interview guide GP-SES-ACC
### Version 1.1, 10-10-2024

This interview guide describes qualitative research using semi-structured interviews for the study "Accessibility and perceived quality of GP care for socioeconomically disadvantaged patients: A qualitative patient-centered interview study".

The duration of the interviews will be approximately 10-20 minutes.

Participants are approached with the question of whether they want to participate in a face-to-face interview following their contact with the general practice.

**Objective**
The aim of these interviews is to gain insight into how patients reach the practice and make an appointment, what problems they may experience in doing so, and whether they have suggestions for improvements. In addition, the interview explores how these patients experience the quality of GP care and what their expectations are.

**Preparation**
Discuss before starting the interview:
- Introduction of interviewer and explanation about study/research goal
- Emphasize that the interview is voluntary and the participant may stop at any time
- Ask permission to record the audio

Turn on audio recording in Teams and start by mentioning the date and participant number.

"We will process and analyze the answers anonymously so that they cannot be traced back to you. We therefore record the interview. This is also done anonymously. There are no right or wrong answers in the interview. You may indicate at any time that you want to stop the interview."

General tips for the interview:
- Ask about what lies behind the participant's answers. "Can you tell me more about that?", "In what respect?", "How come?", "Can you give an example of that?"
- Put yourself as much as possible in the participant's situation so that the conversation can maintain a natural character.
- Ask open questions
- Do not ask questions in a leading way
- No personal opinion/judgment

**Questions**
1. General information
    a. Age, highest completed education, work situation, country of birth, country of birth father/mother
2. Experiences with visiting the GP
    a. Can you talk about your experiences with visiting this GP?
    b. What types of contact have you had with the GP (face-to-face, telephone, e-consultation, etc.)?
    c. How often do you visit the GP?
3. Barriers when making an appointment
    a. a. Have you encountered problems when making an appointment? If so, which ones?
4. Accessibility of the practice
    a. How easy or difficult do you find it to reach the GP when you need an appointment?
    b. Are there certain circumstances that make it more difficult to reach the GP?
    c. Can you describe a situation where you were very satisfied or dissatisfied with the accessibility?
5. Recommendations for improvement
    a. What do you think can be done to improve the accessibility of the GP? Are there specific suggestions you would like to make to improve your experience?
6. Expectations of GP care
    a. What is good GP care for you? What expectations do you have? What do you value?

7. Satisfaction with provided care
    a. How do you find the care here?
    b. What do you think is going well? And why?
    c. Are there things you are dissatisfied with? Things that could be better? Why are you dissatisfied with those?
8. Closing
    a. Are there any other things in this area that you would like to share/say?

"Thank you very much for your cooperation."
