## Supplementary file 2. Interview excerpts for "Accessibility and perceived quality of GP care for socioeconomically disadvantaged patients: a qualitative patient-centered interview study"

| Category | Main theme | Sub-theme | Sub-sub-theme | Sub-sub-sub-theme | Definition | Quote 1 | Quote 2 | Quote 3 |
|---|---|---|---|---|---|---|---|---|
| | **Accessibility** | | | | | | | |
| | | Methods of contacting the practice | | | Different methods patients can use to contact the practice. | | | |
| | | | Walk-in | | The ability to physically go to the practice and make an appointment on the spot. | And, so I prefer, just uh. I live nearby, so if I can just, I come here to the desk to make an appointment. I always walk. | But, you can always drive there, within 10 minutes you're at the GP so you can still make an appointment. | |
| | | | Phone | | The patient calls the practice to ask questions or schedule an appointment. | I usually just call. | I usually call. The GP I currently have is only there on Monday and Tuesday. | Yes, I prefer just by phone or face-to-face and I don't need that app. |
| | | | | Quick phone answering | The practice's ability to answer phone calls quickly. | Well, here I've had a maximum of 2 or 3 so far. And even then it went very quickly. | They answered quickly when I made the appointment to come here. | Yes! I'm almost never on the waiting list, maybe 4 minutes or 5 minutes and then you're helped. So I find that nice. |
| | | | | Long wait for phone to be answered | The time patients have to wait before their call is answered. | Sometimes I have to wait very long, or um, the phone service was down sometimes. | And by phone it's still, I have to wait 20 minutes again for an answer, or 25 minutes. Yes, that doesn't make me happy either. | Yes, I find that convenient to be honest because at other practices you just had to call and then you were just on the phone for 20-30 minutes. Yes, that doesn't really help. |
| | | | | For urgent matters | | For urgent matters if I have an urgent question, for example, or if they don't answer on that app or something, I do call. | | |
| | | | | Efficient/immediate answer | | Yes, it's just press and you get someone on the line. And I'm not a star with the computer, you know. | But it's not direct. You know, by phone is um, and I understand very well that you can't just get a doctor on the phone if you want to ask something. | And when you call, then you have that person on the phone and then you get an answer immediately. So I prefer to just call directly myself. |
| | | | JansApp | | An online app used by the practice to ask questions digitally. | | | |
| | | | | Limited possibilities | Limitations in the functions and options the app offers. | Look, it would be nice if you could also order your medication and so on via that app. | Really just the information, what they give. About the practice. I can find that back on the app. I find that nice, but well you can also find that on the website I think. So that's not such an added value for me yet. But like yes, making that appointment and requesting repeat prescriptions not. | You have um, when you log in you only have 2 options. That is in treatment and completed. And it doesn't matter in which category you ask your question, the treatment is the same. So I don't quite understand what that, where the difference is there. But for the rest you can't put anything in it, so that app should just be expanded in my opinion. |
| | | | | Quick answer to question | The efficiency of receiving answers via the app. | Yes, then I have to enter my code. And then it says you can ask a question. Well, I fill that in and then within well, I always send it early, but I always have an answer within fifteen minutes if the practice is open. | I have used it a few times, especially in the beginning when I had a cold. I've been having a cold for a very long time. And um, yes you do get a good answer to it. And clear too. | Then you don't have to call and you don't have to wait long either. |
| | | | | Long wait for response | The delay patients experience in responses to their questions via the app. | Via the app I have done once, only that is often still waiting a long time for an answer. It says within 30 minutes, but that's not going to work. Not 30 minutes. Sometimes yes, but sometimes also 2 hours or so or 3 hours. Well I think that depends on the question you ask. Because otherwise you would get an answer sooner. | Also an hour later or so, an hour and a half later. Yes, so that was waiting for a while, but I did get an answer to that. | No, um, sometimes I ask a question and then I only get an answer an hour later. And maybe I just want to have a direct answer at that moment. |
| | | | | 24/7 ability to ask questions | The ability to ask questions via the app at any time. | You can just immediately when you have a complaint for example you can immediately text, and then they can help you further with that. | I find that easier, if you're at work and something is wrong or you need to switch, then you can easily send a message like I would like an appointment, is that possible? | |
| | | | | Suitable for small/non-urgent questions | Effective for simple questions that don't require immediate medical attention. | So if I have a short question, I would ask it through there. Then I wouldn't immediately make an appointment I think. | I have um, what did I report now, what was that now, oh yes I have swollen feet. And a few weeks ago on the weekend my feet were so terribly swollen that I thought they were going to burst, and I also took pictures of that. Whole streaks in my leg from my socks, yes I found that quite worrying and I then played that through via the app. | But if I have a small question then I would do that via that chat. |
| | | | | Inefficient/Complicated | The experience that using the app is difficult or impractical. | Yes, I've tried that sometimes, but that is, I don't like it you know. Because you ask a question, and then you get, you have to wait 5 minutes before you get an answer and then you get another question. That keeps going back and forth. | Only I find the app nothing. You can also do repeat prescriptions on the app, but I can't find it back on it. | Because also a lot of things online, things sometimes go wrong. And then you have to do that via such a chatbox, well that doesn't work at all. Because then you always get wrong answers. For example with DHL, then you have to keep going on and after fifteen minutes you finally get someone on the chat. I find that nothing at all, but that's my personal opinion. |
|  |  |  | Email |  | Communication via email for questions or appointments. | Yes I have sent emails, but only for cancelling appointments. |  |  |
| | | Satisfaction | | | | | | |
| | | | Easily accessible/good location | | Accessibility of the practice in terms of location and reachability. | Advantage for me, it's 5 minutes from my house. | It's also easily accessible. I just got off at Zuidplein, and then walked here. | Easily accessible, that for sure. Yes, actually easily accessible. |
| | | | Quick appointment availability | | The speed with which patients can get an appointment. | Not even a few days, maybe 2 days, or 1 day. It really goes very fast yes. | Yes, so far I have. I happened to come last time, when someone here went berserk. They were late, so couldn't be helped anymore and had to wait a week. Or well a week, it was the end of the week but okay. They went berserk. I don't have that, I don't recognize that. | Yes, within a week for sure, within 5 days yes. And then I'm talking about when the weekend is in between, huh. |
| | | | Pharmacy in practice | | Presence of a pharmacy within the healthcare practice for easy access to medication. | And yes, the pharmacy, everything is in one. | And everything runs smoothly with the pharmacy. | Advantage I find is that the pharmacy is already directly in it here, I really find that a plus. |
| | | | Clear website | | A user-friendly and informative website that helps patients find information. | But when I saw this GP practice, I found the site very beautiful and very clearly indicated how you can register and everything. | | |
| | | | Clear when to call | | It is clear to patients at what times they can contact the practice. | I can really always call, in the morning or the afternoon. | They are just always reachable, and they also say in this time we have a break, so after that you can call or text again. | They are just always reachable. |
| | | Dissatisfaction | | | | | | |
| | | | Long wait for repeat prescription | | Delay in obtaining repeat prescriptions. | If I call for a repeat prescription for example then I don't get an answer right away,... or it's not like okay, dude it's good. Because then they look in the computer, done. No, then you get an answer of we have to forward it to the doctor and then, well half a day later you get an answer and if it goes wrong you don't get an answer and you only have your medicines the next day or the day after. So I find that link to getting it sometimes a bit too long myself. | | |
| | | | Difficult to reach | | Difficulties patients experience in contacting the practice. | There it was difficult, because my GP was quite a small GP. Sometimes they don't work, sometimes they're not there. | Very long waiting times. In Hoofddorp, where I was, was really long waiting times and often then it was just indicated on the phone that we'll call you back at another time. Then you don't have to wait that long either. But yes, if you want to arrange something right away then you can't. | Well, I just think, you should put that in a tape. For all you know people come all the way from somewhere here, and then they come all the way here. And then you're just sitting there eating. Look, and if they call then, they don't get to hear well we're at one and we're open again at two o'clock. For example. Or from twelve to two we're closed. |
| | | | Long wait time for appointment | | Frustration due to long waiting times for appointments. | At our old GP you had to wait 3 or 4 weeks before you could get an appointment. Or you could go to a walk-in consultation, but that was always completely full. | Well there's still quite a gap sometimes of 2 weeks. This appointment I have now, that was because a gap had come up where I could jump in between, and otherwise I would have just had to wait 14 days. | At other GPs you have to wait 2 weeks or so before you have an appointment. |
| | | | Strict triage/high threshold for appointment | | The fact that patients cannot just get an appointment, but based on triage. | If I don't feel good then I call and then I say I want to come by now because I really just don't feel good, and pressure in my head and so on. And then they say yes, just take it easy, you already have medication. But I don't feel good at that moment. | They really go very deep into their questions, of how you, and it's already unpleasant if you don't feel good in your skin and you actually just want to go to the GP. You really don't feel like asking all those questions. | There yes, because you really had to be very sick to make an appointment. |

| | | | | | | | |
|---|---|---|---|---|---|---|---|
| | Use of digital technologies | | | | | | |
| | | This era | | The necessity and relevance of digital communication in current healthcare. | Yes, that's the new time huh. You didn't have that before. | Yes, I find it, that's what I like about the modern of now. Personally I like that. | Less. Because this world is becoming more and more digital now so I think it's only good to have such a live chat as well. |
| | | Online consultations | | The possibility to hold consultations via digital channels. | Yes I did have that with my old GP, they did call in the corona time. Then you couldn't go there, only if it was serious, and then you got him via whatsapp. | No, yes, that was in corona time for a while. That they just call, and then you had each other on the phone. | |
| | | | Suitable for certain matters | Effective for specific medical situations or questions. | Yes, look, if it's not really very important to be actually seen I find that can be okay. | But I think, that also depends I think on what you come for. Like yes, look I have your result, then they can see that. | For a result I don't find that such a problem |
| | | | Unsuitable for diagnosis | Limitations of online consultations in making medical diagnoses. | Only, I am of the opinion that you can't really make a diagnosis over the phone. Or with photos. | Yes, photos, yes I mean. You can see someone's skin via the photos but you can't really see well what's really going on. | But if you of course have something wrong and need to be dedicated to an examination that doesn't need to be by phone for me. Because on the phone you can't see what I have in my groin. |
| | | | Impersonal | The experience that digital consultations feel less personal than face-to-face conversations. | I prefer just contact with someone. It's easy and this and that, but yes. Still a bit impersonal I find. | I find that a bit, um, I'm straight right away. Done. And not via online. | I need to hear that person, I need to hear the voice, I just need to see it. That it's just a human, not via a TV or a phone. |
| | | Preference for face-to-face contact | | The preference for personal interaction in medical appointments. | I also find it nicer if it's just face-to-face. | Well, I prefer to just talk to someone. | Yes, I might be very old-fashioned, I prefer just face-to-face. |
| | | | Physical examination possible | The ability to perform physical examinations during a personal consultation. | And I just think that if I'm here with the doctor, then she can maybe check me quicker or whatever. That can't be done over the phone. | Yes they can also check you, and that can immediately also uh, yes. | For a result I don't find that such a problem, but if you of course have something wrong and need to be dedicated to an examination that doesn't need to be by phone for me. Because on the phone you can't see what I have in my groin. |
| | Suggestions for improving | | | Ideas or recommendations from patients to improve accessibility. | I don't know if you have a separate number on the phone line for people who want to make an appointment. If that's not there yet, that might be an idea, that you separate people who want to make an appointment and come into a different waiting queue than people who want to indicate something else for example. | Only, I just think that if you call, then take a tape. Look, every GP who says we're closed until one or until two, you know. And you don't have that here. Then you can just call what you want. | Yes, yes. I have a few times, if you pick up that phone and you're going to call, and then you get that line on the phone. Maybe it's my phone, but if I'm not careful it blasts my head, uh, my ears off my trunk. |
| **Quality of care** | | | | | | | |
| | Satisfaction | | | | | | |
| | | Multiple GPs in practice | | Availability of different GPs for patients. | And here it's nice that there are several. | Yes, I've been to three GPs so far, and all three are just really good. | |
| | | Nice practice | | Aesthetic and physical layout of the practice. | I just find it a really neat practice. | Also from the photos I saw that it was a nice practice, I found that very important too. | It's all good. It's clean here, you can get coffee, you can get tea, you can get everything. |
| | | Speed of service | | The speed with which patients are helped. | And last time I was sick and then I called uh, yes then I couldn't come because I got dizzy and then they would come at 12 o'clock to my home and then they were already there at 11 o'clock so that's positive. | On my appointment, when I came here? Very fast, really waited 2-3 minutes I think. I also came exactly at the time the appointment was. | That's just on time right. I had an appointment with her at half past 10, and I was called in at half pas 10. |
| | | Expertise of the GP | | The knowledge and skills of general practitioners. | I have a rare muscle disease, and yes, he knew a lot about it. | And she knows all about it. Well, when you know that and can immediately provide an answer—because she gave me the same answer I found when I Googled it myself—yeah, I find that impressive. That she can just pull that out of her sleeve like that. | |
| | | Empathy and approachability of the GP | | The attention and understanding shown by general practitioners toward patients. | The way they treat you, and the fact that they simply ask how you're doing. And, yeah, it's not just like patient and doctor. It's more like they take a broader view and ask how things are going, and I really like that. | The communication is also very good, very respectful. | But, when you talk to such a doctor, when they take the time for it, it gives you a really good feeling as a patient. You know, then you leave with a happy feeling. |
| | | Solution-oriented approach | | The GP's focus on finding effective solutions for problems. | And they're very eager to find solutions right away. | Like yesterday, I went, and she still checked if there were ways to help further. So, Dr. *** said she would call my lung specialist to see if anything else could be done, so yeah. | I could get a shot for my back pain right away here, which wasn't possible with my old GP. And now it's solved. |
| | | Young doctors | | The presence of relatively new or young general practitioners. | All young people. Where we came from, they were a bit older, of course. So yeah, I kind of felt like they're still learning, you know. Maybe it gives a more reassuring feeling. | Yeah, I think that's cool! Because they're getting chances, chances to show what they can do. In my time, that wasn't the case. Back then, you had to work harder to prove yourself. But I think it's nice to see young people, I just told her the same. | |
| | | Seeing a regular GP | | The ability to have a long-term relationship with the same GP. | Yeah, because he already knows the story. That just makes it easier. | Yeah, I have that here too. Because you have a practice with three doctors, and I notice that certain doctors take specific patients under their care. You know, like each doctor has, for instance, 30 fixed patients. And I can tell that's the case here. | I think it's nice. She can see what's been discussed, and I go here every month for what's going on with my brain. |
| | | Qualitative treatment | | The quality and effectiveness of the medical care patients receive. | But since I registered here, I've been really well cared for. | And yeah, the care you provide is just good. | And also, what I noticed, I had an appointment the other day, but I forgot about it because I was a bit stressed, and then she called herself saying, 'Hi, you had an appointment with me.' So, I think that's very neat, yeah. And then I could still bring in my urine sample, so I thought that was, yeah, nice. |
| | | Feeling welcome/in place | | The feeling of comfort and acceptance in the practice. | I've never felt so at ease at a GP's practice. | So yeah, she gave such a welcoming feeling. | I'm always warmly received. |
| | | Listening skills of GPs | | The ability of GPs to listen attentively to patients. | They listen carefully to you. | They listen well. | Well, they just help me properly. As I mentioned, they listen. |
| | | Calm waiting room | | The atmosphere and environment of the waiting room that influences patients' experience. | I'm usually alone in the waiting room, and that's something you're not used to. | There's a calming atmosphere here. And that's really a plus. Also, I have a son with autism, so it's quiet here. And that, yeah, I think that's something more practices should do. Other GPs could learn from this. | I find it pretty quiet here in the waiting room, but maybe it's because we're used to it being super crowded at the other one. |
| | | Clear explanations and communication | | The GP's ability to communicate and explain clearly. | They explain things well here. For instance, the last time when my daughter had her tonsils, the GP said she's going to grow out of it, it'll pass, you know, they really reassure you. | And my doctor also gave me quite a bit of information about my health. | For example, like now, I also get additional information via email that I can read through if I didn't understand something. |
| | | Attention to the patient | | The focus of GPs on the individual needs of patients. | More attention to the patient, for someone who comes with a problem. | I came here for the first time, and I wanted to get my blood tested because four weeks ago I had been on medication from the other GP, and I just wanted to know how my blood was doing after four weeks of medication. So, when I told her I wanted to have my blood tested, she said, 'But that's already been done.' I said, 'But I want to know how my blood is now, after four weeks of medication.' So she did take the blood test. | But when you talk to a doctor, if they take the time for you, it really gives you a great feeling as a patient. You know, then you leave with a happy feeling. |
| | | Modern practice | | The contemporary facilities and technology in the practice. | I love a more modern practice, and I think you are one. Personally, I like the use of modern tools and thinking. I'm not into the old-fashioned way. | And then when I came here, yeah, I really like the modern approach, the way you think about things, I like that. | |
| | | Timely referral | | The promptness and effectiveness of referrals to specialists or further care. | I get referred immediately if needed. | But after I came here, I was immediately referred to the hospital, and it turned out I had gallstones in my gallbladder. So, yeah, I'm happy I now know where the pain comes from. | She helped me then. She said, 'I still want you to go to the hospital.' Oh yeah, for my lungs, that's what it was. |
| | Dissatisfaction | | | | | | |
| | | Unpleasant behavior | | Unfriendly or inappropriate behavior from healthcare providers. | And if it wasn't really necessary, there was still some sharpness in the response, which I think is wrong. | | |
| | | No answer to questions | | The feeling that questions are not taken seriously or are not answered. | Then, yeah, I actually didn't get an answer anywhere. | And also, when I've had tests done, like blood or urine tests because of my slightly elevated kidney levels and high blood pressure, they don't call back. | Not getting answers. |
| | | Looking things up during consultation | | Situations where doctors search for information during the consultation rather than having it prepared in advance. | And then they spent a lot of time looking at the computer. | His knowledge, he looks up a lot on the internet. | My old doctor used to Google things. Well, when that happens, you know you're not well-informed. You haven't studied enough. |
| | | Uncomfortable feeling | | General discomfort or dissatisfaction with the experience in the practice. | He just didn't make me feel comfortable. | It was really unpleasant. I had to have something burned off here, and when I was lying on the examination table, I thought, you know, they could just put that wireless burner or sensor on the table. Why does it have to go here (points to pocket)? You know, from then on I thought, I'm out of here. I just got a really bad feeling from him. | |

| | | | | | |
|---|---|---|---|---|---|
| | No regular GP | The absence of a consistent GP, leading to inconsistency in care. | First of all, it was a very large practice. It's at ***, where I think there used to be about 16 doctors. And now, lately, there have only been three, and they only work half-days. And yeah, that just doesn't work. Every time I wanted to make an appointment, I had to tell the whole story because I kept getting a different doctor. And yeah, I didn't want that anymore. | When you come here, you get different doctors all the time. I prefer to have just one GP who knows my history. | Well, here I've already seen 5 or 6 different GPs since we've been coming here. And it doesn't matter, I mean, they know what they're talking about, and they help me. But you can't build a relationship with them. |
| | Not hear/believed | The feeling that the patient's concerns or complaints are not taken seriously. | With my old GP, it was more like, you know, when my little daughter kept getting sick with her tonsils. I'd go straight to the GP to see what's going on, because sometimes she gets short of breath. And my old GP would say, 'Yeah, you come too often for this.' But if my daughter is sick, I have no other choice. I try the first three days on my own, and if it doesn't get better, I just go. That was really frustrating. | And he kept giving me prednisone for the inflammation, which made me swell up. And if you said anything, he just made up his own stories. | Well, not really, because I had a complaint, and there was never any examination done. And the complaint just kept coming back. |
| | Different perspective | A gap between the GP's experiences/expectations and those of the patient. | Or they'd just say, 'You can do this or that,' and that was it. But I have quite a few things wrong with me, so I prefer it when they actually look into it. | Sometimes they find your concerns tedious, or if you have a problem, they say it's not that serious, that you can just take paracetamol. But you think, 'No, that's not enough, you know.' | When I'm not feeling well, I call and say, 'I really need to come in, I don't feel well, I have pressure in my head.' And they say, 'Take it easy, you already have medication.' But at that moment, I don't feel well. |
| Expectations of the GP | | | | | |
| | Clear communication and explanation | The need for clear and understandable information from the GP. | Clear language, so that I understand what is being said. | | |
| | Trusting relationship | The relationship between the GP and patient, based on trust and respect. | And for example, that you can just comfortably tell your complaints, what you are struggling with, and so on. | You just want to build a strong rapport with a GP. That's important. | Yes, the GP is a trusted person. And I just think it's important that when I tell you something that's supposed to be confidential, as a doctor, I don't want that shared with another doctor. Or another one after that. Because then it's not a secret anymore. The more people who know, the less I can rely on it. |
| | Shared-decision making | The process where the GP and patient make decisions together about treatment. | And also being asked what you expect, what you would like. | Let's say a GP might have one point of view, but I have mine too. If I feel much worse than I did a few weeks ago, then I expect that if I ask to have my blood checked, that it should just be done. | Well, for example, if something is wrong, and then, what they think you should do about it. For instance, medications or treatments, you know. A patient has their own opinion too, and it should be emphasized. It can work for or against you, that's true. But generally, yes, I tend to speak up if I think differently. |
| | Knowledge and expertise | The expertise and information the GP has. | That the doctor immediately understands you. Yes, I've had a few appointments with her, and no matter what I bring up, she just knows exactly what's going on. | They examine everything properly. | They are, of course, medically trained and know much more than I do. I'm just a simple cleaner. |
| | Attention and empathy | The GP's focus on the emotional and physical needs of the patient. | The most important thing for me is that they listen carefully to you. | That the doctor listens to you, listens to the complaint, and helps you with it. | That they listen to you, that they give you attention. |
| | Getting an appointment quickly | The ability to get an appointment quickly without long waiting times. | Whether people could schedule an appointment at short notice. I often saw that they couldn't book an appointment or the practice was just closed. Yes, then you think, I don't want to go there. Because healthcare is important, of course. | And I also think that if there is a real emergency, you should be able to get in quickly. | And sometimes I really need a doctor quickly. Because yes, I had a double pulmonary embolism 10 years ago, so I was resuscitated 3 times. So I find it reassuring to be able to see a doctor right away. |
| | Solution for the problem | The GP's ability to provide effective solutions to the patient's concerns. | And also offering a solution for what I'm going to the GP for. | If I'm satisfied with what the doctor says, or what can be done, and gives me advice. | Looking for solutions for your health. |
| | Good accessibility | The GP's availability for questions or appointments. | Definitely accessibility. That's number one. | Actually, whether it's located nearby. | |
| | Feeling comfortable | The feeling that patients feel welcome and safe at the practice. | I have my own health issues, my son's, and my partner's, so you want to feel comfortable where you are. | That you also feel at home here, you know. | |
| | Timely referral | The promptness with which a GP refers patients to specialists or further care. | Or to where I should be referred. | And if they don't trust something themselves, they send you to the hospital. | Or refer you. |
